# Challenges and proposed solutions in making clinical research on COVID-19 ethical. A status quo analysis across German research ethics committees

**DOI:** 10.1101/2020.08.11.20168773

**Authors:** Alice Faust, Anna Sierawska, Katharina Krüger, Anne Wisgalla, Joerg Hasford, Daniel Strech

## Abstract

**Background:** In the course of the COVID-19 pandemic, the biomedical research community’s attempt to focus the attention on fighting COVID-19, led to several challenges within the field of research ethics. However, we know little about the practical relevance of these challenges for Research Ethics Committees (RECs).

**Methods:** We conducted a qualitative survey across all 52 German RECs on the challenges and potential solutions with reviewing proposals for COVID-19 studies. We de-identified the answers and applied thematic text analysis for the extraction and synthesis of challenges and potential solutions that we grouped under established principles for clinical research ethics.

**Results:** We received an overall response rate of 42%. The 22 responding RECs reported that they had assessed a total of 441 study proposals on COVID-19 until 21 April 2020. For the review of these proposals the RECs indicated a broad spectrum of challenges regarding i) social value (e.g. lack of coordination), ii) scientific validity (e.g. provisional study planning), iii) favourable risk-benefit ratio (e.g. difficult benefit assessment), iv) informed consent (e.g. strict isolation measures), v) independent review (e.g. lack of time), vi) fair selection of trial participants (e.g. inclusion of vulnerable groups), and vii) respect for study participants (e.g. data security). Mentioned solutions ranged from improved local/national coordination, over guidance on modified consent procedures, to priority setting across clinical studies.

**Conclusions:** RECs are facing a broad spectrum of pressing challenges in reviewing COVID-19 studies. Some challenges for consent procedures are well known from research in intensive care settings but are further aggravated by infection measures. Other challenges such as reviewing several clinical studies at the same time that potentially compete for the recruitment of in-house COVID-19 patients are unique to the current situation. For some of the challenges the proposed solutions in our survey could relatively easy be translated into practice. Others need further conceptual and empirical research. Our findings together with the increasing body of literature on COVID-19 research ethics, and further stakeholder engagement should inform the development of hands-on guidance for researchers, funders, RECs, and further oversight bodies.

## Background

In December 2019, an outbreak of the previously unknown coronavirus SARS-CoV-2 that likely occurred in Hubei Province in China drew the world’s attention. Subsequently, the virus spread rapidly on a global scale and led the WHO to declare a pandemic emergency on 11 March 2020. With the sudden outbreak of the novel virus resulting in COVID-19 disease, the international biomedical research community aimed to better understand the virus and disease and engaged in the development of therapies, diagnostics and prevention measures. On 27 June 2020, the registry clinicaltrials.gov listed 2,341 clinical studies, of which 1,314 (56%) were classified as interventional studies, and 257 clinical trials on COVID-19 were listed in the EudraCT database of the European Medicines Agency (EMA). The true number of clinical studies is probably much higher because most health-related observational studies are not prospectively registered [1].

The rapidly growing number of studies of one disease at the same time raises concerns about research ethics and best practices. Can clinical research that is planned, funded, reviewed, conducted and published in a very short time fulfil the necessary requirements of effective, efficient, and ethical science? To support the research community in these unprecedented times, the WHO published the document “Ethical standards for research during public health emergencies: Distilling existing guidance to support COVID-19 R&D” on 29 March 2020 [2], which refers to already existing recommendations for ethical research during pandemics and briefly summarizes important points. The “Guidance on the management of clinical trials during the COVID-19 pandemic” of the European Commission and the EMA outlines some specific recommendations, for example, on informed consent [3]. Expert papers point out ethically relevant risks and the potential damage caused by poorly planned and conducted research and stress the importance of adhering to scientific standards in times of crisis [4].

These extraordinary pandemic circumstances most likely also pose challenges for the research ethics committees (RECs) that are in charge of the assessment of COVID-19 studies. However, there is little information available on what challenges RECs currently face and how they deal with those challenges. To the knowledge of the authors, only one report exists that describes which types of modifications were necessary in 41 reviewed proposals and explanatory documents reviewed at one Chinese hospital [5].

The objective of this study was to broaden the understanding of current challenges in the work of RECs through a status quo analysis across all German RECs.

## Methods

Sampling: The sample included 52 German RECs that participate in the assessment of clinical study proposals as required by German law and professional regulations and that are members of the umbrella organization “Association of Medical Ethics Committees in Germany” (AKEK: Arbeitskreis Medizinischer Ethik-Kommissionen).

Questionnaire: To study the “qualitative spectrum of challenges and proposed solutions” in the most efficient way, we developed a questionnaire with three open questions. In addition, RECs were asked to indicate the number of interventional studies and non-interventional study proposals assessed until 21 April 2020. For further details, see the original questionnaire in the appendix.

Survey: The survey was conducted between 21 April 2020 and 30 April 2020. The questionnaire was sent by e-mail together with a cover letter from the AKEK office, and the responses were returned to the AKEK office. The anonymized questionnaires were forwarded to the involved investigators (AF, AS, and DS) of the QUEST Center for analysis.

Analysis: To extract, analyse, and synthesize the relevant information on the challenges and proposed solutions mentioned in the responses from the 22 RECs, thematic text analysis was performed independently by two researchers (AF, AS) using MaxQDA version 2020. First, the codes were grouped under one or more principles as described in an internationally established framework for clinical research ethics [6]. Second, response passages mentioning challenges and solutions were identified, and descriptive codes were applied. Third, the coding results were compared to identify potential differences in coding. However, only minor differences occurred, which were solved through discussion. Fourth, themes mentioned in one response were matched with those from another response to collate the various codes and cluster the findings into categories and subcategories of challenges and solutions. All researchers discussed and slightly modified the matrix for internal consistency and agreed on the final matrix.

## Results

A questionnaire was sent to 52 RECs, of which 22 (42%) participated in the survey. According to information from the AKEK office, these 22 RECs together assessed 50% of the total 15,501 study proposals in Germany in 2017 and 53% of the total 17,182 study proposals in 2018.

The 22 RECs reported that they had assessed a total of 441 study proposals on COVID-19 as of 21 April 2020. These proposals included 229 proposals for interventional COVID-19 studies, of which 42 related to German drug law (AMG: Arzneimittelgesetz), one related to German medical device law (MPG: Medizinproduktgesetz) and 187 related to the German professional code for physicians (Berufsrecht). In addition, there were 212 proposals for non-interventional studies.

The qualitative responses from the 22 RECs on perceived challenges and proposed solutions were all grouped under one or more of seven principles of the employed research ethics framework: social value, scientific validity, informed consent, respect for participants, independent review, favourable risk-benefit analysis and fair participant selection. We did not identify any responses that could be grouped under the eigths principle collaborative partnership. The analysis reached thematic saturation at the framework level. Thematic saturation implies that no new principles or other overarching themes, but only further subcategories, could be generated. Table 1 presents all challenges and proposed solutions derived from the thematic text analysis. In the following, we explain selected topics that were addressed more frequently or with diverse viewpoints in narrative form.

**Table 1:** Qualitative spectrum of challenges and potential solutions in the review of COVID-19 trial proposals by research ethics committees (RECs)

| Ethical principles | Challenges<br>(sub-categories with examples of original responses) | Potential solutions<br>(sub-categories with examples of original responses) |
| --- | --- | --- |
| Social value | <u>Lack of coordination/structures in study planning</u><br>• Frequent simultaneous planning of several COVID-19 studies within an individual hospital and uncoordinated submission of studies to the local REC<br>• Large number of small studies that partly overlap thematically and are often poorly structured<br>• Lack of concrete objectives | <u>Coordination</u><br>• A multitude of studies could be coordinated at the institutional level.<br><u>Priority setting</u><br>• The urgency of studies in the current pandemic situation should be balanced. Several projects could be carried out retrospectively. More detailed assessment of the expected knowledge gains should be performed. |
| Scientific validity | <u>Insufficient information for RECs</u><br>• Scarce or incomplete proposals or study protocols<br>• Frequent lack of biometrics<br>• Insufficient disclosure of the funding<br>• Lack of detailed description of data management or the consent process (for non-AMG/MPG* studies)<br>• Difficulty evaluating valid/relevant endpoints when the knowledge of the disease is still uncertain, leading to difficulties in calculating sample sizes<br><u>Poor, provisional study planning/lack of clear objectives/insufficient statistics</u><br>• Study protocols with the character of a rough sketch | <u>Better information</u><br>• Requests from experts (e.g., for suggested biometrics) and additional REC meetings to discuss proposals in detail could be adopted.<br>• Studies could be subdivided into one main study with the possibility of submitting (ethically more problematic) sub-studies via amendments.<br>• If the essential aspects of the trial are evident from the proposal documents and there are no fundamental concerns, a positive vote with comments is given, indicating which aspects still must be supplemented in the study protocol.<br>• Pragmatic assessment of initial proposal could be tolerated if an interim evaluation were planned.<br><u>Improvement of study planning and advisory process</u><br>• Documents could be prepared with the help of institutional |
|  | • Lack of clear rationale<br>• Lack of documentation of independent factors influencing the outcome of the study<br>• Insufficient number of planned study participants<br>• Questionable study design selected for proof of efficacy<br>• Poorly prepared and elaborated surveys/registry studies<br>• Unclear length of the study (e.g., patients should be included as long as the pandemic lasts)<br>• Frequent requirement of revisions/changes to primary and secondary endpoints<br>• Lack of clarity regarding whether the proposal reflects a clinical study or experimental treatment according to national law ("individueller Heilversuch")<br>• Provision of statistical advice in advance rarely possible<br>• In interventional studies, frequent inadequacy of sample size calculations (e.g., initially only small sample sizes, which are then continuously increased)<br>• Lack of applicant knowledge of which guidelines they should follow | core facilities for clinical studies.<br>• Telephone/online advice could be offered<br>• In REC counselling, a focus could be placed on scientific aspects, and (if possible) proposal formalities could be reduced.<br>• A statistician could be involved; an analysis plan and clear research objectives could be required.<br>• Additionally, the REC chair could contact principal investigator/sponsor for quick clarification of questions/problems.<br>• COVID-19-specific assistance with the application process could be offered.<br>• The REC should contact statistician before voting.<br>• If there are too few COVID-19 patients locally, multicentre studies with a sufficient number of patients could be required. |
| Informed consent | <u>Informed consent for patients unable to give consent</u><br>• For intensive care patients, a lack of an informed consent process, with the reasoning that these patients for the study are unable to give consent (according to national law: § 9 para. 2 GDPR)<br>• Lack of clarity regarding how patients can be included in clinical studies when no patient-representative is appointed or available<br>• Acceptance of verbal consent by some studies<br>• Inability to give consent, even among non-intubated patients, due to the severity of the disease<br>• Unclear whether additional, explicit consent for taking biospecimens should be required<br>• Processing of consent via telephone without a description | <u>Alternative forms of consent</u><br>• Patients unable to give informed consent may be presented with the documents after they have gained the ability to give consent.<br>• Consent can be given by legal representatives; additionally, subsequent signatures of the legal representatives by post or fax should be considered.<br>• More detailed description in the study protocol of the process of including patients who are unable to give consent could be provided.<br>• For pure data collection where only clinical routine data are to be collected for secondary use, there is no time pressure. In this case, it is possible to wait until the patient is either able to give consent again or a legal |
|  | of documentation and verifiability<br>• Problems obtaining consent from caregivers if they are not allowed to come to the hospital<br>• Restricted possibility of consulting/informing relatives due infection control measures<br>• Lack of permission for consent document to leave the patient's room due to infection control measures<br><u>Formal problems with consent</u><br>• Partial unavailability of patient information<br>• Lack of comprehensibility, unclear wording,<br>• Lack of information about data protection<br>• Too many AMG/MPG* amendments; difficulties ensuring that the content is checked | representative who can give consent has been identified. The data can then be entered retrospectively as soon as consent has been obtained. If the patient were to die in the meantime, the data could be recorded anonymously.<br>• In the case of deceased persons, samples could be required to be anonymous, and no identifying research (whole or partial genome sequencing) would be carried out.<br>• A photo of the signed consent form can be archived outside the patient room.<br><u>Formal improvement</u><br>• Comprehensibility could be improved, and information about data protection should be added.<br>• REC should provide support and advice on all consent-related points.<br>• In addition to individual REC advice, European and national recommendations are given to sponsor/applicant.<br>• The necessary clarification documents could be archived.<br>• Sometimes, re-submission should also be required.<br>• The collection of data, samples, images, etc., could be enabled in crisis situations without consent according to national Infection Protection Law. |
| Respect for participants |  | <u>Improved data management</u><br>• Secondary use of only anonymized data at the end of the usually relatively short course of the disease would also make informed consent unnecessary.<br>• Regarding biobanks, the proposed project could be restricted to COVID-19 and infectious diseases. |
|  |  | Additionally, consent to anonymization after the end of the study/use for non-specified research projects should be obtained. |
| Independent review | <u>Lack of time/time pressure</u><br>• Co-counselling of the locally responsible REC in case of significantly shortened deadlines for COVID-19 studies<br>• Applicants' expectation of rapid feedback<br>• Intensive workload<br>• Lack of courses/training for PIs due to infection protection measures<br>• Frequent emergency staffing of RECs | <u>Better time management</u><br>• Those reviews of proposals on non-COVID-19 topics could be postponed that are required by the National code for physicians.<br>• In multicentre studies, after several unsuccessful attempts to contact other responsible RECs, assessment should be performed without their involvement.<br>• In multicentre-studies, additional claims from RECs responsible for secondary research sites are formulated as a suspensive condition.<br>• Consultation outside the usual deadlines and additional meetings could be conducted while maintaining the usual quality of consultation.<br>• Prioritization of proposals could be adopted.<br>• With regard to the investigators' courses, more tolerance might be necessary, with at least temporary acceptance of online courses.<br>• Communication with the REC members could be performed via a web-based cloud instead of regular physical jour fixes.<br>• Communication with applicants could be performed via e-mail and short-term review by chairperson.<br>• Non-interventional clinical studies could be reviewed with secondary importance. Most of their questions can also be approached slightly later. |
| Favourable risk-benefit ratio | <u>Uncertainty about the disease</u><br>• In the case of interventional trials, difficulty regarding uncertainty about the disease because there is little preliminary data available or the | <u>Reduction in the amount/frequency of blood samples</u><br>• For blood tests, consent to use residual blood from routine examination could be obtained (after the patient regains the ability to give consent). |
|  | situation is constantly changing, leading to a necessity for a certain degree of extrapolation<br>• Difficulty of assessment due to the heterogeneous clinical picture<br>• Limited benefit assessment due to the currently insufficient knowledge about the "natural course" of the disease<br>• Amount/frequency of blood samples insufficiently justified |  |
| Fair participant selection | <u>Inclusion and allocation criteria</u><br>• Unclear rationales for the exclusion of certain COVID-19 patient groups<br>• Planning of several clinical studies despite the limited availability of hospitalized COVID-19 patients: How to allocate patients? Based on what criteria?<br><u>Vulnerable populations</u><br>• Increasing number of studies exclusively with clinical and nursing staff | <u>Protection of vulnerable populations</u><br>• Statements on the careful handling of particularly vulnerable occupational groups in this critical situation could be made. |

With regard to **scientific validity**, some RECs complained about a lack of relevant information to assess the study validity or pointed to inadequate statistics. Comments also highlighted the partial lack of a clear rationale for “repurposing studies”. Some RECs mentioned that it was apparent from the proposals that the applicants were under time pressure and that this pressure partly negatively affected the methodological quality of the submissions. The mentioned solutions to these challenges were diverse and in part contradictory. Some respondents tolerated a “pragmatic” assessment of the submitted documents. Others preferred additional meetings to discuss challenging issues in-depth. The requirement of biometric advice before the submission COVID-19 applications was mentioned as a strategy to ensure an effective and efficient advisory process and enable applicants to plan their studies better.

On the topic of **informed consent**, the vulnerability of patients requiring intensive care and facing isolation as an infection control measure was highlighted as a particular challenge. The isolation of COVID-19 patients makes direct contact with caregivers/legal proxy decision makers difficult. Many study proposals aimed explicitly or implicitly to include patients who were unable to give or restricted from giving informed consent. In addition, the RECs seemed to be uncertain or insufficiently prepared with regard to guidance on alternative or modified consent formats. A further problem arose from the question of which groups of COVID-19 patients were to be classified as unable to give or restricted from giving consent and according to which criteria. Suggested solutions for the inclusion of persons unable to give informed consent were to collect consent by proxy and/or deferred consent. The importance of written consent was noted; however, the possibility of consent by telephone and the use of photographs of the original documents in isolation situations were proposed as solutions as well.

The **social value** principle was challenged by the conduct of several insufficiently coordinated and thematically difficult-to-distinguish COVID-19 studies in one hospital/region. REC members also highlighted a general lack of clear target actions in the planning of several register projects. The coordination of studies at the university level or at the national level was mentioned as a possible solution. Another suggestion was an explicit priority setting for research projects.

Many RECs reported intensive time pressure in the processing of COVID-19 proposals that we identified as a challenge for the **independent review** principle. Due to the lack of time, RECs reported difficulties in guaranteeing a high-quality assessment of all submitted proposals. In addition, RECs mentioned a strong demand of the applicants for a quick assessment. Logistical problems, such as working from the home office, would make things even more difficult. The solutions proposed included additional REC meetings, prioritized assessment of certain types of proposals and the use of online services for communication within the REC and with applicants. Some RECs mentioned the option to focus their assessments on proposals for which their institutions hosted the lead principal investigator and to fast-track multicentre proposals for which their institutions only served as a cooperating research facility.

The principle of the **fair selection of study participants** was challenged, for example, by the frequent inclusion of clinical staff in studies. Furthermore, RECs struggled with participant selection because the number of required study participants exceeded the number of available COVID-19 patients. It was unclear for RECs how to allocate patients across studies or how to determine and rank “priority studies”. As a proposed solution for the protection of hospital staff, RECs recommended that the applicants provide statements “on the careful handling of particularly vulnerable hospital staff”. No potential solutions were mentioned for the allocation/priority setting problem.

A **favourable risk-benefit ratio** was difficult to pursue due to insufficient knowledge about COVID-19 and its heterogenic and rapidly changing clinical picture. The problems arose especially in intervention studies. Above all, the benefits for the participating patients were difficult to assess. Regarding risk, for example, the use of non-therapeutic research procedures, such as increased frequency of blood sampling, was partly insufficiently justified. A solution to this problem could be to check whether residual blood from routine care could be used. No solutions were mentioned for the problem of the difficulty of assessing benefits.

Finally, RECs reported various challenges with data management and data protection regarding the sensitive information of study participants, which we assigned to the principle of **respect for study participants**. Proposed solutions included more anonymization or pseudonymization of data and a limitation of the use of the data to COVID-19 specific research projects.

## Discussion

In a national survey of all 52 German RECs from April 2020, we studied the number of assessed COVID-19 study proposals and the qualitative spectrum of associated challenges and proposed solutions. The 22 RECs reported that they assessed 441 COVID-19 study proposals (229 interventional and 212 non-interventional). The reported challenges and proposed solutions were grouped under eight research ethics principles [6].

In the following, we supplement the survey results described above with a more detailed interpretation and information on initiatives that have been started since April to directly address some of the challenges in the coming months.

Shortly after the survey was distributed, the German Ministry of Education and Research (Bundesministerium für Bildung und Forschung: BMBF) funded the National Research Network, which, under the direction of Charité-Universitätsmedizin Berlin, is working on various approaches to improve the efficiency and effectiveness of national research on COVID-19 [7]. This network has the potential to strengthen **scientific validity** by providing, for example, standardized data sets for COVID-19 projects, a national database and measures for the coordination and creation of quality standards in medical research on COVID-19. In addition to harmonization and coordination, expert contributions highlighted the importance of not lowering the bar regarding the scientific validity of individual COVID-19 studies [4].

Regarding **informed consent** for persons unable to give consent or with restricted ability to give consent, there are already various previous experiences and recommendations in the context of emergency and intensive care medicine on the topic [8-10]. To address this topic in a practice-oriented way for COVID-19 research, these recommendations should be further developed and specified for isolated patients, acknowledging several infection control measures. COVID-19-specific recommendations on the topic of deferred consent or the monitoring of consent processes (“consent monitor”) might be of particular relevance [11, 12].

There is little previous experience with forecasting the **social value** of individual clinical studies [13], especially with priority setting across clinical studies in a pandemic situation. The prioritization of research projects is usually addressed from a long-term perspective and focuses on the prioritization of whole research areas [14]. For short-term prioritization, it might be possible to agree on ethically relevant prioritization criteria such as “clinical relevance” and a “sufficiently high probability of success”. While the general clinical relevance of various therapeutic approaches to COVID-19 might be determined relatively well, there are important challenges in clarifying the likelihood of their success. The error rate of early clinical research is generally very high [15], and there is a lack of robust concepts for identifying study projects with a particularly high probability of success.

The exploratory survey reported here has the following limitations. First, many responses focused on the areas of scientific validity and informed consent, which may be related to the fact that the questionnaire explicitly asked about challenges and solutions regarding “statistics/study quality”, “informed consent”, and “other issues”. However, the broad spectrum of challenges and proposed solutions mentioned shows that many responding RECs expanded the focus. Second, we received a response rate of 42%. It is possible that the RECs that responded were the RECs where particular challenges in connection with COVID-19 studies frequently arose. As described above, our survey did not aim to make a statement about the frequency of challenges but rather about the qualitative spectrum of the challenges described. Third, we could not verify the information on the number of applications processed.

Currently, there are many international contributions that address ethical issues in COVID-19 research, such as in “challenge studies” [16] or in “high-demand trials” [17]. Our status quo analysis on ethical issues based on feedback from 22 German RECs broadens our understanding of the spectrum of ethical challenges in COVID-19 research as perceived from those involved in the concrete review and oversight of COVID-19 studies. Further research on ethical challenges and proposed solutions as perceived by principle investigators and other stakeholder groups could complement this picture. Practice-oriented recommendations for the most pressing ethical challenges should be developed to support applicants, RECs, funders, potential research participants, and proxy decision makers in the best possible way (“pandemic response”) and to prepare for future pandemic situations (“pandemic preparedness”). The BMBF-funded project “PRECOPE - Preparedness and Response for Ethical Challenges in Human Subject Research during COVID-19 and similar PandEmics”, starting in August 2020, will address these tasks. Based on a systematic literature review, in-depth interviews, and further stakeholder engagement, PRECOPE aims to develop practice-oriented recommendations for the most pressing ethical challenges. As most ethical challenges in COVID-19 research ethics are expected to be on a global scale, international cooperation in developing preparedness and response measures is of utmost importance.

## Data Availability

The datasets used and/or analysed during the current study may be available from the corresponding author on reasonable request. There are legal constraints that prohibit us from making all data publicly available, e.g., they are containing information that could compromise the de-identification of the participating RECs.

## Declarations

## Acknowledgements

We would like to thank the participating RECs for their time and willingness to cooperate.

## Ethics approval and consent to participate

Not applicable.

## Author’s contributions

DS and JH participated in the conceptualization of the status-quo analysis. JH, KK and AW collected the data. AF and AS analyzed and interpreted the data. AF and DS wrote the manuscript. AS, JH, AW, KK provided feedback on the manuscript. All authors read and approved the final manuscript.

